# Cutaneous immune-related adverse events are associated with longer overall survival in advanced cancer patients on immune checkpoint inhibitors: a multi-institutional cohort study

**DOI:** 10.1101/2023.01.16.23284635

**Authors:** Shijia Zhang, Kimberly Tang, Guihong Wan, Nga Nguyen, Chenyue Lu, Pearl Ugwu-Dike, Neel Raval, Jayhyun Seo, Nora A. Alexander, Ruple Jairath, Jordan Phillipps, Bonnie W. Leung, Kathleen Roster, Wenxin Chen, Leyre Zubiri, Genevieve Boland, Steven T. Chen, Hensin Tsao, Shadmehr Demehri, Nicole R. LeBoeuf, Kerry L. Reynolds, Kun-Hsing Yu, Alexander Gusev, Shawn G. Kwatra, Yevgeniy R. Semenov

## Abstract

**Background:** Cutaneous immune-related adverse events (cirAEs) occur in up to 40% of immune checkpoint inhibitor (ICI) recipients. However, the association of cirAEs with survival remains unclear.

**Objective:** To investigate the association of cirAEs with survival among ICI recipients.

**Methods:** ICI recipients were identified from the Mass General Brigham healthcare system (MGB) and Dana-Farber Cancer Institute (DFCI). Patient charts were reviewed for cirAE development within 2 years after ICI initiation. Multivariate time-varying Cox proportional hazards models, adjusted for age, sex, race/ethnicity, Charlson Comorbidity Index, ICI type, cancer type, and year of ICI initiation were utilized to investigate the impact of cirAE development on overall survival.

**Results:** Of the 3,731 ICI recipients, 18.1% developed a cirAE. 6-month landmark analysis and time-varying Cox proportional hazards models demonstrated that patients who developed cirAEs were associated with decreased mortality (HR=0.87,p=0.027), particularly in melanoma patients (HR=0.67,p=0.003). Among individual morphologies, lichenoid eruption (HR=0.51,p<0.001), psoriasiform eruption (HR=0.52,p=0.005), vitiligo (HR=0.29,p=0.007), isolated pruritus without visible manifestation of rash (HR=0.71,p=0.007), acneiform eruption (HR =0.34,p=0.025), and non-specific rash (HR=0.68, p<0.001) were significantly associated with better survival after multiple comparisons adjustment.

**Limitations:** Retrospective design; single geography.

**Conclusion:** CirAE development is associated with improved survival among ICI recipients, especially melanoma patients.

**Capsule Summary:**

- Patients on immune checkpoint inhibitors (ICIs) who developed cutaneous immune-related adverse events (cirAEs) had favorable outcomes. This was especially notable for melanoma patients who had cirAEs, both those with vitiligo and other morphologies.
- Development of cirAEs in ICI-treated patients can be used to prognosticate survival and guide treatment decisions.

## Introduction

Immune checkpoint inhibitors (ICIs) have transformed cancer care over the past decade. Checkpoint proteins are membrane-bound markers that bind T-cell partner proteins to induce anergy^1^. ICIs block these interactions, strengthening endogenous immune responses against tumors. Currently-approved therapies target Programmed cell death protein 1 (PD-1) and its ligand (PD-L1) or Cytotoxic T-lymphocyte protein 4 (CTLA-4)^1^. As of 2022, 9 ICIs were approved and with expansion to different targets and cancers, the percentage of cancer patients eligible for ICIs has increased from 1.54% in 2011 to 43.6% in 2020^2,3^.

Immune activity alterations from ICI can manifest as immune-related adverse events (irAEs), necessitating informed management guidelines^4^. Most common and least lethal irAEs affect the skin and gastrointestinal tract, whereas rarer but more serious irAEs affect the cardiovascular and central nervous systems. Notably, cutaneous irAEs (cirAEs), the most common toxicities, occur in up to 40% of recipients and can be potential low-grade biomarkers of therapeutic effect^5^.

Though early reports have associated irAE development with improved ICI response, studies examining individual contributions of cirAEs to predicting outcomes are limited to single-institutional observational studies with small sample size and absence of appropriate control populations^6,7^. A recent study from our group utilized population-level claims-based databases to investigate outcomes among patients with cirAEs, identifying significantly increased survival^8^. However, this was limited by diagnostic uncertainty in cirAE definitions. Therefore, in this study we examine overall and morphology-specific survival implications of cirAE development among ICI recipients using a large-scale multi-institutional clinical registry with manual cirAE phenotyping. We seek to provide necessary confirmatory data on the clinical implications of these toxicities, which will inform treatment decisions and patient counseling.

## Methods

### Cohort Determination

Cancer patients receiving ICI at Massachusetts General Hospital, Brigham and Women’s Hospital, and Dana-Farber Cancer Institute between December 1, 2011 and October 30, 2020 were included. Patients were excluded if records were incomplete for variables of interest. A flowchart is in **Figure 1**.

**Figure 1.**
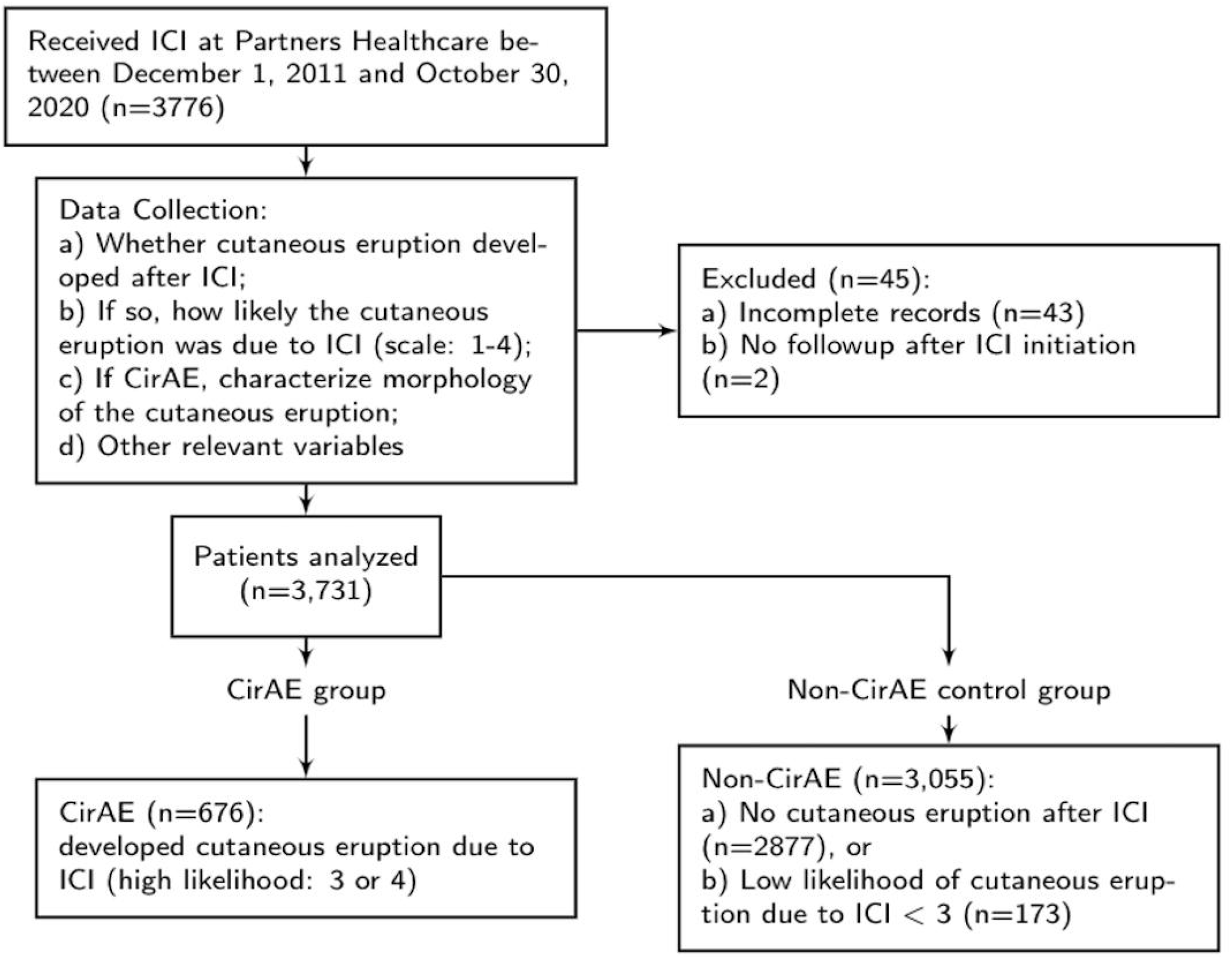
Study Design Flowchart Attached as separate jpeg file in accordance with the JAAD submission checklist

### Data Extraction

Research Patient Data Registry (RPDR)^9^ and Enterprise Data Warehouse (EDW) are institutional clinical databases in the Mass General Brigham (MGB) Healthcare System. RPDR contains multimodal clinical information: basic demographics, International Classification of Diseases (ICD) codes, and lab results. EDW provides detailed documentation of all ICI administrations.

Patient age, sex, race/ethnicity, cancer type, ICD codes, date of death or last follow-up were extracted. Due to small sample sizes among some races/ethnicities, this variable was categorized into White, Asian, Black, (non-white) Hispanic, and other. ICD codes from all visits were used to calculate Charlson Comorbidity Index (CCI) score at ICI initiation^10^. EDW was used to ascertain time and class of ICI received. ICIs were separated into anti-PD-1 (pembrolizumab, nivolumab, cemiplimab), anti-PD-L1 (atezolizumab, avelumab, durvalumab), anti-CTLA-4 (ipilimumab), and combination therapy (CTLA-4 and either PD-1 or PD-L1 inhibition).

### Chart Review

Manual chart review was conducted by two independent reviewers to ascertain cirAE presence, timing, and morphology following ICI initiation. Suspected events were categorized and graded using Common Terminology Criteria for Adverse Events version 5.0^11^. For each patient who developed a cutaneous eruption following ICI, a likelihood score was assigned on a scale of 1 (highly unlikely) to 4 (highly likely). The likelihood of cirAE was determined by review of rash timing, morphology, absence of competing risk factors, histologic confirmation when available, and response to treatment of the eruption. Patients who achieved likelihood scores of 3 and 4 were categorized as having a cirAE. If concordance was not achieved, a third reviewer arbitrated the cases. Further details are provided in **Supplemental Methods**.

### Statistical Analyses

We used Pearson’s chi-squared or Fisher exact test for categorical variables and t-test or Kruskal-Wallis test for continuous variables. Kaplan-Meier curves were utilized to investigate differences in survival between patients with and without cirAEs. For patients with no known mortality information, their last system encounters were their censoring dates. We performed landmark survival analyses and time-varying Cox proportional hazards regression modeling, adjusting for age, sex, race/ethnicity, CCI, ICI type, cancer type, and year of ICI initiation to account for guarantee-time bias^12^. Guarantee-time bias occurs when exposure of interest competes with risk of mortality (e.g., patients must be alive long enough to be able to develop a cirAE). Not controlling for this would bias the model in favor of longer survival in the cirAE cohort. To account for this, we performed a landmark analysis, assessing exposure (e.g., cirAE status) before the given landmark time and excluding patients with the outcome of interest (e.g., mortality) prior to the landmark^13^. A survival model was then utilized on this restricted population to evaluate the relationship between exposure and outcome. Sensitivity analyses were performed around landmark times: 3-, 6-, 9-, and 12-months following ICI initiation. Another control for guarantee-time bias is utilizing time-varying Cox proportional hazards regression modeling, which uses the exposure as a time-dependent covariate; subjects are classified as unexposed until the start of cirAE and exposed thereafter^14^. This model has the advantage of incorporating all study follow-up data since the analysis begins at the time of cohort entry. Therefore, this method has increased statistical power over the landmark method^15^. Lastly, because all patients in this cohort had metastatic cancer, the CCI was rescaled to exclude cancer diagnoses. The analyses were repeated for individual cancer types and cirAE morphologies. All statistical analyses were conducted in R statistical software version 3.6.1.

## Results

A total of 3,731 ICI recipients were identified. Patient characteristics are shown in **Table 1**. Between cirAE and non-cirAE groups, there were no significant differences in median age at ICI initiation (65.1 vs 65.4 years), mean CCI score (3.30 vs 3.64), and sex (42.9% vs 46.0% female). Compared to the non-cirAE group, the cirAE group had longer duration of follow-up (913 vs 385 days), higher proportion of white patients (93.8% vs 90.4%), higher proportion of combination immunotherapy utilization (17.8 % vs 8.5 %), and higher proportion of melanoma patients (35.7% vs 17.1%), all which were statistically significant. **Table 2** demonstrates the distribution of cirAE morphologies among all cancer, melanoma, and non-melanoma patients.

**Table 1.**
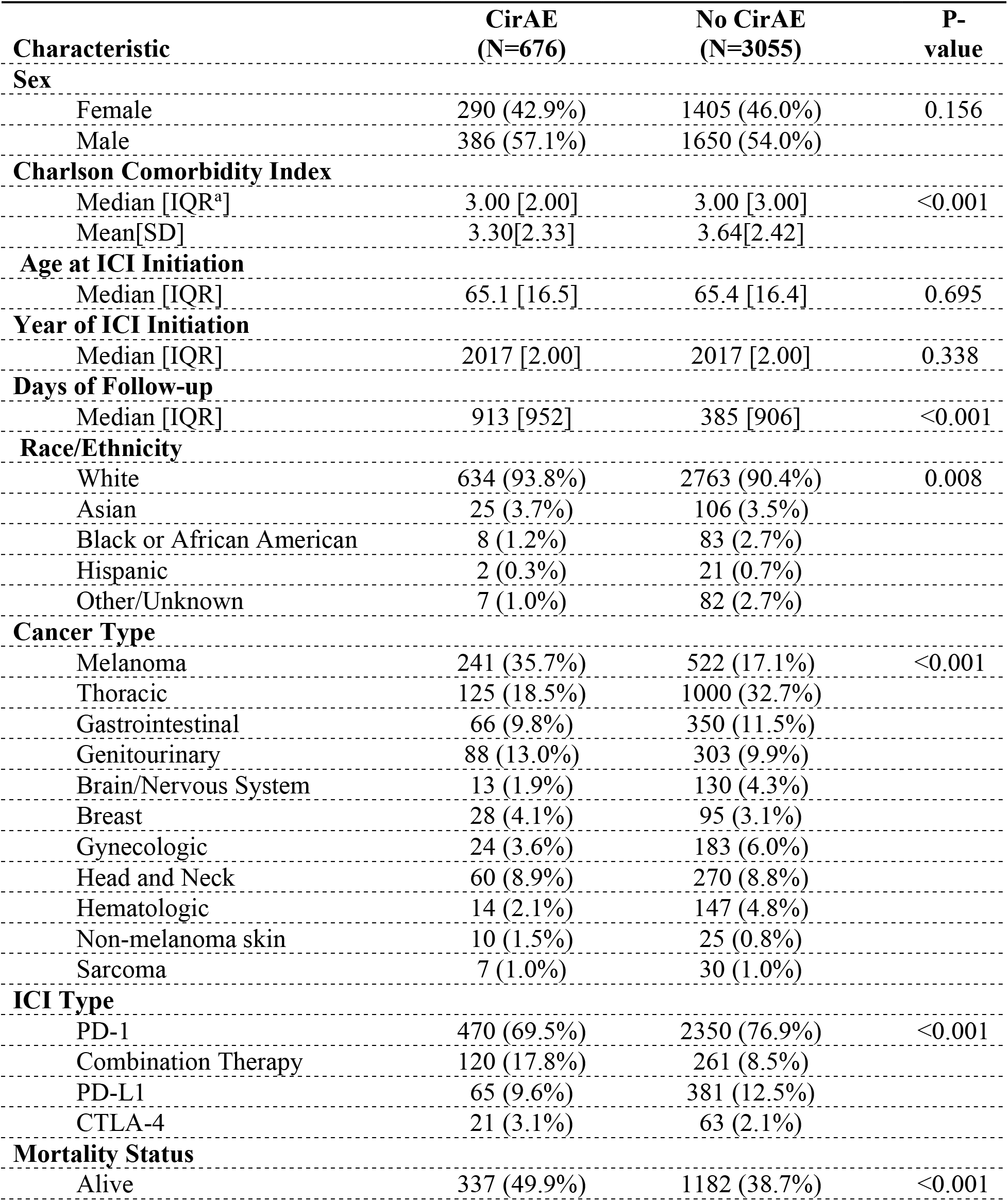
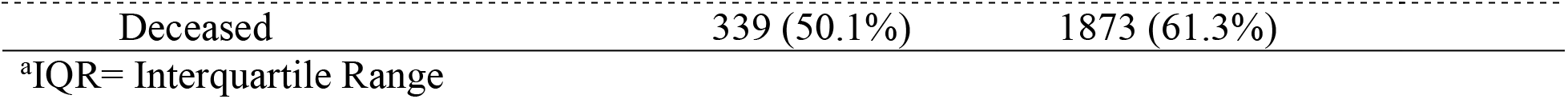
Baseline Characteristics for Cancer Patients Treated with ICI

| **Characteristic** | **CirAE**<br>**(N=676)** | **No CirAE**<br>**(N=3055)** | **P-**<br>**value** |
| --- | --- | --- | --- |
| **Sex** |  |  |  |
| Female | 290 (42.9%) | 1405 (46.0%) | 0.156 |
| Male | 386 (57.1%) | 1650 (54.0%) |  |
| **Charlson Comorbidity Index** |  |  |  |
| Median [IQR<sup>a</sup>] | 3.00 [2.00] | 3.00 [3.00] | <0.001 |
| Mean[SD] | 3.30[2.33] | 3.64[2.42] |  |
| **Age at ICI Initiation** |  |  |  |
| Median [IQR] | 65.1 [16.5] | 65.4 [16.4] | 0.695 |
| **Year of ICI Initiation** |  |  |  |
| Median [IQR] | 2017 [2.00] | 2017 [2.00] | 0.338 |
| **Days of Follow-up** |  |  |  |
| Median [IQR] | 913 [952] | 385 [906] | <0.001 |
| **Race/Ethnicity** |  |  |  |
| White | 634 (93.8%) | 2763 (90.4%) | 0.008 |
| Asian | 25 (3.7%) | 106 (3.5%) |  |
| Black or African American | 8 (1.2%) | 83 (2.7%) |  |
| Hispanic | 2 (0.3%) | 21 (0.7%) |  |
| Other/Unknown | 7 (1.0%) | 82 (2.7%) |  |
| **Cancer Type** |  |  |  |
| Melanoma | 241 (35.7%) | 522 (17.1%) | <0.001 |
| Thoracic | 125 (18.5%) | 1000 (32.7%) |  |
| Gastrointestinal | 66 (9.8%) | 350 (11.5%) |  |
| Genitourinary | 88 (13.0%) | 303 (9.9%) |  |
| Brain/Nervous System | 13 (1.9%) | 130 (4.3%) |  |
| Breast | 28 (4.1%) | 95 (3.1%) |  |
| Gynecologic | 24 (3.6%) | 183 (6.0%) |  |
| Head and Neck | 60 (8.9%) | 270 (8.8%) |  |
| Hematologic | 14 (2.1%) | 147 (4.8%) |  |
| Non-melanoma skin | 10 (1.5%) | 25 (0.8%) |  |
| Sarcoma | 7 (1.0%) | 30 (1.0%) |  |
| **ICI Type** |  |  |  |
| PD-1 | 470 (69.5%) | 2350 (76.9%) | <0.001 |
| Combination Therapy | 120 (17.8%) | 261 (8.5%) |  |
| PD-L1 | 65 (9.6%) | 381 (12.5%) |  |
| CTLA-4 | 21 (3.1%) | 63 (2.1%) |  |
| **Mortality Status** |  |  |  |
| Alive | 337 (49.9%) | 1182 (38.7%) | <0.001 |

|  |  |  |
| --- | --- | --- |
| Deceased | 339 (50.1%) | 1873 (61.3%) |

<sup>a</sup>IQR= Interquartile Range

**Table 2:** Distribution of CirAE Morphologies in All Cancer, Melanoma, and Other Cancer Patients

| Cutaneous Diagnosis | All Cancer<br>(N=676) | Melanoma<br>(N=241) | Other Cancer*<br>(N=435) | P-value |
| --- | --- | --- | --- | --- |
| Acneiform Eruption | 13 (1.9%) | 5 (2.1%) | 8 (1.8%) |  |
| Bullous Eruption | 17 (2.5%) | 6 (2.5%) | 11 (2.5%) |  |
| Drug Hypersensitivity<br>NOS | 32 (4.7%) | 14 (5.8%) | 18 (4.1%) |  |
| Eczematous Eruption | 41 (6.1%) | 11 (4.6%) | 30 (6.9%) |  |
| Erythema-multiforme-<br>like Eruption | 4 (0.6%) | 2 (0.8%) | 2 (0.5%) |  |
| Isolated Pruritus without<br>visible manifestation of<br>rash | 90 (13.3%) | 35 (14.6%) | 55 (12.6%) |  |
| Lichenoid Eruption | 52 (7.7%) | 10 (4.1%) | 42 (9.7%) | <0.001 |
| Maculopapular Eruption | 121 (17.9%) | 56 (23.2%) | 65 (14.9%) |  |
| Mucositis | 9 (1.3%) | 1 (0.4%) | 8 (1.8%) |  |
| Panniculitis | 3 (0.4%) | 1 (0.4%) | 2 (0.5%) |  |
| Psoriasiform Eruption | 30 (4.4%) | 8 (3.3%) | 22 (5.1%) |  |
| Rash NOS | 222 (32.8%) | 74 (30.8%) | 148 (34.0%) |  |
| SJS/TEN-like Eruption | 4 (0.6%) | 1 (0.4%) | 3 (0.7%) |  |
| Vitiligo | 22 (3.3%) | 17 (7.1%) | 5 (1.1%) |  |
| Alopecia | 1 (0.1%) | 0 (0%) | 1 (0.2%) |  |
| DRESS | 1 (0.1%) | 0 (0%) | 1 (0.2%) |  |
| Follicular Eruption | 5 (0.7%) | 0 (0%) | 5 (1.1%) |  |
| Sweet's Syndrome | 3 (0.4%) | 0 (0%) | 3 (0.7%) |  |
| Urticarial Eruption | 6 (0.9%) | 0 (0%) | 6 (1.4%) |  |

\*Other cancer includes: thoracic, gastrointestinal, genitourinary, brain/nervous system, breast, gynecologic, head and neck, hematologic, non-melanoma skin cancer, and sarcoma.

Multivariate time-varying Cox proportional hazards models (**Table 3**) demonstrated that patients with cirAEs were significantly associated with better survival when analyzed across all cancer indications (HR=0.87, p=0.027). No differences in mortality by race/ethnicity or sex were noted. Independent of cirAE development, patients with gastrointestinal malignancies (HR=1.32, p<0.001) had worse outcomes than thoracic cancer patients, while patients with melanoma (HR=0.44, p<0.001) and genitourinary cancer (HR=0.79, p=0.002) had better survival. To explore the contribution of ICI-induced vitiligo, a further analysis categorized patients into those who developed vitiligo and non-vitiligo cirAEs (**eFigures 1a and 1b**), demonstrating that both groups were significantly associated with decreased mortality compared to patients without cirAEs among all ICI recipients and melanoma patients. On average, patients who developed cirAEs survived 320 days (10.5 months) longer than those who did not.

**Table 3:** Association between CirAE with Overall Survival Using a Multivariate Time-varying Cox Proportional Hazards Model

| **Characteristic** | **Hazard Ratio** | **95% Confidence Interval** | **P-value** |
| --- | --- | --- | --- |
| **CirAE** | 0.87 | 0.79, 0.98 | 0.027 |
| **Sex** |  |  |  |
| Female | Ref<sup>a</sup> | Ref |  |
| Male | 1.04 | 0.95, 1.13 | 0.4 |
| **Age at ICI Initiation** | 1.00 | 0.99, 1.00 | 0.4 |
| **Charlson Comorbidity Index** | 1.09 | 1.07, 1.11 | <0.001 |
| **Race/Ethnicity** |  |  |  |
| White | Ref | Ref |  |
| Asian | 1.08 | 0.86, 1.36 | 0.5 |
| Black | 0.90 | 0.69, 1.18 | 0.4 |
| Hispanic | 1.02 | 0.60, 1.74 | >0.9 |
| Other/Unknown | 0.84 | 0.62, 1.14 | 0.3 |
| **Cancer Type** |  |  |  |
| Thoracic | Ref | Ref |  |
| Melanoma | 0.44 | 0.38, 0.51 | <0.001 |
| Gastrointestinal | 1.32 | 1.15, 1.51 | <0.001 |
| Genitourinary | 0.79 | 0.68, 0.92 | 0.002 |
| Other* | 0.94 | 0.84, 1.05 | 0.3 |
| **ICI Type** |  |  |  |
| CTLA-4 | Ref | Ref |  |
| Combination Therapy | 1.22 | 0.88, 1.69 | 0.2 |
| PD-1 | 0.94 | 0.69, 1.27 | 0.7 |
| PD-L1 | 0.77 | 0.55, 1.09 | 0.12 |
| **Year of ICI Initiation** | 1.05 | 1.01, 1.09 | 0.014 |

<sup>a</sup>Ref= Reference group
\*Other includes: brain/nervous system, breast, gynecologic, head and neck, hematologic, non-melanoma skin cancer, and sarcoma

To explore the association of cirAEs with each cancer type, we performed multivariate time-varying Cox proportional hazards models restricting the population to patients with a specific malignancy (**eTables 1**). Here, association of cirAEs with good prognosis was strongest among melanoma patients (HR=0.67, p=0.003) (**eTable 2)**. CirAEs were not significantly associated with mortality when restricting to thoracic, gastrointestinal, genitourinary, head and neck, and other cancers individually. When grouping all non-melanoma cancers, there was a weak association that did not reach statistical significance (**eTable 3**).

These analyses were repeated using a landmark approach. A 6-month landmark was utilized for the primary model as most cirAEs occurred by this time (**eTable 4)**^15^. Sensitivity analyses were performed using 3-, 9-, and 12-months as alternative landmark times (**eTable 5-8)**. As before, patients who developed cirAEs demonstrated significant survival advantage using landmark times of 3 months (HR=0.75,p<0.001) and 6 months (HR=0.83,p=0.006), with better survival at 9 months (HR=0.89,p=0.11) and 12 months (HR=0.94,p=0.5) which did not reach statistical significance. Additionally, we conducted the same landmark and sensitivity analyses among melanoma patients, as this is where the association of cirAEs was strongest in the time-varying Cox models (**eTable 9-12**). Among these patients, cirAE development was associated with significantly lower risk of mortality at 3 months (HR=0.65,p=0.002), 6 months (HR=0.65,p=0.003), 9 months (HR=0.64,p=0.004), and 1 year (HR=0.68,p=0.021) as landmark times.

We next investigated the adjusted survival impact of individual cirAE morphologies (**Table 4)**. Compared with the non-cirAE group, almost all morphologies were associated with better survival. Specifically, rash (HR=0.68, p<0.001), lichenoid eruption (HR=0.51,p<0.001), psoriasiform eruption (HR=0.52,p =0.005), vitiligo (HR=0.29,p=0.007), isolated pruritus without visible manifestations of rash (HR=0.71,p=0.007), and acneiform eruption(HR=0.34,p=0.025) remained significantly associated with better survival after Benjamini-Hochberg (BH) correction (**eTables 13-19)**. Eczematous eruption (HR = 0.71, p=0.056) was borderline significantly associated.

**Table 4:** Association of CirAE Morphology with Overall Survival

| **Cutaneous Morphology<sup>a</sup>** | **Hazard Ratio** | **95% Confidence Interval** | **P-value<sup>b</sup>** |
| --- | --- | --- | --- |
| Vitiligo | 0.29 | 0.12, 0.71 | 0.007* |
| Acneiform Eruption | 0.34 | 0.13, 0.87 | 0.025* |
| Lichenoid Eruption | 0.51 | 0.36, 0.73 | <0.001* |
| Psoriasiform Eruption | 0.52 | 0.33, 0.82 | 0.005* |
| Rash NOS | 0.68 | 0.57, 0.81 | <0.001* |
| Isolated Pruritus Without Visible<br>Manifestation of Rash | 0.71 | 0.55, 0.91 | 0.007* |
| Eczematous Eruption | 0.71 | 0.50, 1.01 | 0.056 |
| Drug Hypersensitivity NOS | 0.79 | 0.52, 1.21 | 0.3 |
| Maculopapular Eruption | 0.82 | 0.66, 1.03 | 0.084 |
| Bullous Eruption | 0.87 | 0.45, 1.68 | 0.7 |
| Other CirAE Morphologies | 0.98 | 0.64, 1.50 | >0.9 |

<sup>a</sup>Each row corresponds to a separate multivariable time-varying Cox proportional hazards model for the association of cirAE morphology with overall survival by comparison to patients without cirAEs, adjusting for age at ICI initiation, gender, race, CCI, cancer type, ICI type, and year of ICI initiation.
<sup>b</sup>P-values that remain significant after Benjamini-Hochberg multiple comparison adjustment are marked with an asterisk.

## Discussion

Though irAEs have been shown to be important indicators of ICI efficacy, their association with mortality is a growing field of interest ^16-21^. This large-scale retrospective study investigates the association of cirAEs with mortality outcomes among ICI-treated cancer patients using expert-phenotyped cirAE status. We demonstrate that cirAE development after ICI initiation is an important prognostic indicator of response to ICI, particularly among melanoma patients.

CirAEs occurred in 18% of ICI recipients in contrast to clinical trial reports of cirAEs occurring in up to 40% of patients. This may be from cirAE overdiagnosis in clinical trials, which typically lack rigorous phenotyping. Additionally, cirAEs reported in observational studies are usually symptomatic enough to require clinical evaluation, whereas cirAEs reported in clinical trials may be mild. The latter group is unlikely detected in real-world observational data as patients may not present for evaluation, thus, explaining this discrepancy.

Overall, our study suggests that cirAE development is associated with a 13% reduction in mortality, with patients developing cirAEs living on average 10.5 months longer than those who did not. The association of cirAEs with increased survival identified here is consistent with our prior population-level findings (using algorithmic rather than manual cirAE curation done here), which demonstrated a 19-23% statistically significant risk reduction in mortality among patients with cirAEs^8^. The overall similar conclusions reached by these different analytical approaches with completely independent cohorts provide additional rigor to our study findings. Additionally, almost all cirAE morphologies were associated with better survival, with isolated pruritus without visible manifestation of rash, rash, lichenoid eruption, psoriasiform eruption, acneiform eruption and vitiligo demonstrating the most significant correlation: 29%, 32%, 49%, 48%, 66% and 71% reduction in mortality, respectively. These findings are important as cirAEs are among the earliest events to occur following ICI initiation and could therefore be early predictive biomarkers of therapeutic response.

When stratifying by cancer type, the association of cirAEs with improved survival was strongest in patients with melanoma. Restricting the population to melanoma patients, those who developed cirAEs had a 33% decreased risk of mortality compared to those who did not. We speculate that the survival improvement seen among melanoma patients may be explained by melanoma’s high tumor mutational burden ^22-23^ as previous studies have demonstrated that high levels of immunogenic neoantigens in melanoma is correlated with increased response to anti-PD-1 and better survival^22-23^. This may also trigger reactivity to similar skin antigens, contributing to cirAE development and better outcomes. But for most individual cancers, cirAEs were not significantly correlated. Excluding melanoma and examining all non-melanoma cancers as a group demonstrated weak clinical protection of cirAEs that ceased to remain statistically significant.

Due to the melanoma cohort’s strong signal, we performed subgroup analyses of individual cirAE morphologies among melanoma patients showing that both vitiligo and non-vitiligo cirAEs are associated with significantly longer survival. Though previous studies identified vitiligo as associated with decreased mortality among melanoma patients, we show that other morphologies of cirAEs are also associated with better outcomes in comparison to patients without cirAEs^24-26^. Even after excluding vitiligo, cirAE development remains associated with better survival among all ICI recipients and melanoma patients specifically. Landmark analyses performed yielded similar conclusions, even after sensitivity analyses around landmark time-3-,6-, 9-, and 12-months. Notably, lichenoid eruption demonstrated significant association with increased survival that surpassed that of vitiligo in the setting of melanoma.

A possible explanation for the association of vitiligo development with better survival in melanoma is epitope spreading. This develops when immune responses against primary epitopes spread to other distinct epitopes in the same tissue type^27-30^. This results in clonal diversification of T-cell responses, proving to be beneficial in cancer treatment. When expansion of reactivity occurs toward tumor antigens, it enhances the ICI efficacy^29-30^. We hypothesize that this phenomenon is responsible for development of vitiligo when T-cells to melanoma antigens respond inappropriately to new epitopes on normal melanocytes^27-30^. Because epitope spreading contributes to robust antitumor activity, this may explain the increased survival among ICI recipients with melanoma who develop vitiligo when compared to those who do not, particularly due to expected shared melanocyte antigens.

However, the development of other cirAEs among melanoma patients that confer comparable correlation with better survival than vitiligo, leads us to believe that cirAE development in the setting of melanoma may not be entirely driven by epitope spreading. Studies have suggested that systemic induction of the immune response by ICIs may re-activate pre-existing T-cells, which originally developed against melanocytes at the onset of melanoma^30^.

Systemic memory T-cells can express receptors important in the return of these circulating lymphocytes to the original tissue of activation, ^31-33^ manifesting as cirAEs. This idea of tissue homing suggests that tumor tissue source is an important variable in generation of irAEs, and the interaction between prognostically favorable cirAE morphologies and tumor tissue type may uncover mechanistic insights into immunotherapy response.

This study is limited by retrospective design. Patients may not be representative as they were drawn from two tertiary care cancer centers. Differences in clinical judgment may exist between physicians who documented notes and medically trained researchers who extracted information from charts. To adjust for inter-rater biases, cirAE status was ascertained by two independent reviewers. Cases that did not meet concordance were evaluated by a board-certified dermatologist with expertise in immunotherapy toxicities (YRS). To account for immortal time and selection bias, landmark analyses and time-varying Cox proportional hazards models were used to provide a more rigorous approach.

Although there are studies analyzing the association of irAEs with mortality, we demonstrate significantly favorable outcomes in ICI-treated patients who develop cirAEs, with enhanced protection in melanoma patients. We find cirAEs to be associated with increased survival as a group and additionally, significantly improved prognosis is observed among other morphologies in addition to vitiligo. Together, this suggests that cirAE development after ICI is a key prognostic indicator of survival. This is important in guiding oncologists and dermatologists in managing cirAEs and counseling patients about the prognostic implications of these toxicities. Further studies should investigate interactions between management of toxicities and survival and underlying biologic mechanisms for development of these favorable events and their interaction with immunotherapy response.

## Data Availability

All data produced in the present study are available upon reasonable request to the authors

## Acknowledgements

We thank Stacey Duey from Partners Scientific Computing for her help in extracting RPDR patient data.

## References

1. He X, Xu C. Immune checkpoint signaling and cancer immunotherapy. Cell research. 2020;30(8):660–669.

2. Jamal S, Hudson M, Fifi-Mah A, Ye C. Immune-related adverse events associated with cancer immunotherapy: a review for the practicing rheumatologist. The Journal of rheumatology. 2020;47(2):166–175.

3. Haslam A, Gill J, Prasad V. Estimation of the percentage of US patients with cancer who are eligible for immune checkpoint inhibitor drugs. JAMA network open. 2020;3(3):e200423–e200423.

4. Brahmer JR, Lacchetti C, Schneider BJ, et al. Management of immune-related adverse events in patients treated with immune checkpoint inhibitor therapy: American Society of Clinical Oncology Clinical Practice Guideline. Journal of clinical oncology: official journal of the American Society of Clinical Oncology. 2018;36(17):1714.

5. Thompson LL, Krasnow NA, Chang MS, et al. Patterns of cutaneous and noncutaneous immune-related adverse events among patients with advanced cancer. JAMA dermatology. 2021;157(5):577–582.

6. Hirotsu KE, Scott MKD, Marquez C, et al. Histologic subtype of cutaneous immune-related adverse events predicts overall survival in patients receiving immune checkpoint inhibitors. J Am Acad Dermatol. 2021 Dec 4:S0190-9622(21)02922-4. doi: 10.1016/j.jaad.2021.11.050. Epub ahead of print. PMID: 34875301.

7. Otto TS, Chang MS, Thompson LL, Chen ST. Limitations of morphology-based management for immune checkpoint inhibitor-related cutaneous adverse events. J Am Acad Dermatol. 2021 Jun;84(6):e281–e282. doi: 10.1016/j.jaad.2021.01.054. Epub 2021 Jan 23. PMID: 33493573.

8. Tang K, Seo J, Tiu BC, et al. Association of Cutaneous Immune-Related Adverse Events With Increased Survival in Patients Treated With Anti-Programmed Cell Death 1 and Anti-Programmed Cell Death Ligand 1 Therapy. JAMA Dermatol. 2022 Feb 1;158(2):189–193. doi: 10.1001/jamadermatol.2021.5476. PMID: 35019948; PMCID: PMC8756357.

9. Nalichowski R, Keogh D, Chueh HC, Murphy SN. Calculating the benefits of a Research Patient Data Repository. AMIA. Annual Symposium proceedings AMIA Symposium. Published online 20068.

10. Hall WH, Ramachandran R, Narayan S, Jani AB, Vijayakumar S. An electronic application for rapidly calculating Charlson comorbidity score. BMC Cancer. 2004;4:94. Published 2004 Dec 20. doi:10.1186/1471-2407-4-94

11. “Common Terminology Criteria for Adverse Events (CTCAE).” Cancer Therapy Evaluation Program (CTEP), National Cancer Institute, 21 Sept. 2020, https://ctep.cancer.gov/protocolDevelopment/electronic_applications/ctc.htm.

12. Lévesque LE, Hanley JA, Kezouh A, Suissa S. Problem of immortal time bias in cohort studies: example using statins for preventing progression of diabetes. Bmj. 2010;340.

13. Gleiss A, Oberbauer R, Heinze G. An unjustified benefit: immortal time bias in the analysis of time-dependent events. Transpl Int. 2018 Feb;31(2):125–130. doi: 10.1111/tri.13081. Epub 2017 Nov 9. PMID: 29024071.

14. Zhang Z, Reinikainen J, Adeleke KA, Pieterse ME, Groothuis-Oudshoorn Cgm. Time-varying covariates and coefficients in Cox regression models. Ann Transl Med. 2018;6(7):121. doi:10.21037/atm.2018.02.12

15. Cho, I.S., Chae, Y.R., Kim, J.H. et al. Statistical methods for elimination of guarantee-time bias in cohort studies: a simulation study. BMC Med Res Methodol 17, 126 (2017). https://doi.org/10.1186/s12874-017-0405-6

16. Wongvibulsin S, Pahalyants V, Kalinich M, et al. Epidemiology and risk factors for the development of cutaneous toxicities in patients treated with immune checkpoint inhibitors: A United States population-level analysis. J Am Acad Dermatol. Published online 2021:1-10.

17. Das S, Johnson DB. Immune-related adverse events and anti-tumor efficacy of immune checkpoint inhibitors. J Immunother Cancer. 2019 Nov 15;7(1):306. doi: 10.1186/s40425-019-0805-8. PMID: 31730012; PMCID: PMC6858629.

18. Albandar, Heidar J et al. “Immune-Related Adverse Events (irAE) in Cancer Immune Checkpoint Inhibitors (ICI) and Survival Outcomes Correlation: To Rechallenge or Not?.” Cancers vol. 13,5 989. 27 Feb. 2021, doi:10.3390/cancers13050989

19. Petrelli F, Grizzi G, Ghidini M, et al. Immune-related Adverse Events and Survival in Solid Tumors Treated With Immune Checkpoint Inhibitors: A Systematic Review and Meta-Analysis. Journal of immunotherapy (Hagerstown, Md : 1997). 2020;43(1). doi:10.1097/CJI.0000000000000300

20. Street S, Chute D, Strohbehn I, et al. The positive effect of immune checkpoint inhibitor-induced thyroiditis on overall survival accounting for immortal time bias: a retrospective cohort study of 6596 patients. Annals of Oncology. 2021;32(8). doi:10.1016/j.annonc.2021.05.357

21. Burke KP, Grebinoski S, Sharpe AH, Vignali DAA. Understanding adverse events of immunotherapy: A mechanistic perspective. J Exp Med. 2021 Jan 4;218(1):e20192179. doi: 10.1084/jem.20192179. PMID: 33601411; PMCID: PMC7754677.

22. Strickler JH, Hanks BA, Khasraw M. Tumor Mutational Burden as a Predictor of Immunotherapy Response: Is More Always Better? Clin Cancer Res. 2021 Mar 1;27(5):1236–1241. doi: 10.1158/1078-0432.CCR-20-3054. Epub 2020 Nov 16. PMID: 33199494

23. Kim SI, Cassella CR, Byrne KT. Tumor Burden and Immunotherapy: Impact on Immune Infiltration and Therapeutic Outcomes. Front Immunol. 2021 Feb 1;11:629722. doi: 10.3389/fimmu.2020.629722. PMID: 33597954; PMCID: PMC7882695.

24. Matsuya T, Nakamura Y, Matsushita S, Tanaka R, Teramoto Y, Asami Y, et al. Vitiligo expansion and extent correlate with durable response in anti-programmed death 1 antibody treatment for advanced melanoma: A multi-institutional retrospective study. J Dermatol. 2020 Jun;47(6):629–635. doi: 10.1111/1346-8138.15345. Epub 2020 Apr 10. PMID: 32275100.

25. Nakamura Y, Tanaka R, Asami Y, Teramoto Y, Imamura T, Sato S, et al. Correlation between vitiligo occurrence and clinical benefit in advanced melanoma patients treated with nivolumab: A multi-institutional retrospective study. J Dermatol. 2017 Feb;44(2):117–122. doi: 10.1111/1346-8138.13520. Epub 2016 Aug 11. PMID: 27510892.

26. Teulings HE, Limpens J, Jansen SN, Zwinderman AH, Reitsma JB, Spuls PI, et al. Vitiligo-like depigmentation in patients with stage III-IV melanoma receiving immunotherapy and its association with survival: a systematic review and meta-analysis. J Clin Oncol. 2015 Mar 1;33(7):773–81. doi: 10.1200/JCO.2014.57.4756. Epub 2015 Jan 20. PMID: 25605840.

27. Brossart P. The Role of Antigen Spreading in the Efficacy of Immunotherapies. Clin Cancer Res. 2020 Sep 1;26(17):4442–4447. doi: 10.1158/1078-0432.CCR-20-0305. Epub 2020 May 1. PMID: 32357962.

28. Hardwick N, Chain B. Epitope spreading contributes to effective immunotherapy in metastatic melanoma patients. Immunotherapy. 2011 Jun;3(6):731–3. doi: 10.2217/imt.11.62. PMID: 21668310.

29. Lo JA, Kawakubo M, Juneja VR, Su MY, Erlich TH, LaFleur MW, et al. Epitope spreading toward wild-type melanocyte-lineage antigens rescues suboptimal immune checkpoint blockade responses. Sci Transl Med. 2021 Feb 17;13(581):eabd8636. doi: 10.1126/scitranslmed.abd8636. PMID: 33597266; PMCID: PMC8130008.

30. Chan LS, Vanderlugt CJ, Hashimoto T, Nishikawa T, Zone JJ, Black MM, et al. Epitope spreading: lessons from autoimmune skin diseases. J Invest Dermatol. 1998 Feb;110(2):103–9. doi: 10.1046/j.1523-1747.1998.00107.x. PMID: 9457902.

31. Campbell JJ, Haraldsen G, Pan J, Rottman J, Qin S, Ponath P, et al. The chemokine receptor CCR4 in vascular recognition by cutaneous but not intestinal memory T cells. Nature. 1999 Aug 19;400(6746):776–80. doi: 10.1038/23495. PMID: 10466728.

32. Fuhlbrigge RC, Kieffer JD, Armerding D, Kupper TS. Cutaneous lymphocyte antigen is a specialized form of PSGL-1 expressed on skin-homing T cells. Nature. 1997 Oct 30;389(6654):978–81. doi: 10.1038/40166. PMID: 9353122.

33. Jiang X, Clark RA, Liu L, Wagers AJ, Fuhlbrigge RC, Kupper TS. Skin infection generates non-migratory memory CD8+ T(RM) cells providing global skin immunity. Nature. 2012 Feb 29;483(7388):227–31. doi: 10.1038/nature10851. PMID: 22388819; PMCID: PMC3437663.

34. Chen S, LeBoeuf N, Reynolds K, Semenov Y. 1263 Dermatologic immune related adverse event disease definitions: a multi-institutional Delphi consensus project presented on behalf of the oncodermatology working group. vol 10. 2022:A1309-A1309.

35. Cohen, J. (1960). A Coefficient of Agreement for Nominal Scales. Educational and Psychological Measurement, 20(1), 37–46. https://doi.org/10.1177/001316446002000104

