## Supplement materials for "Cutaneous immune-related adverse events are associated with longer overall survival in advanced cancer patients on immune checkpoint inhibitors: a multi-institutional cohort study"

### Online-only Supplementary Material

#### Supplementary Methods

We defined our patient population by identifying all patients who received ICI therapy from billing records. CirAE phenotyping was completed in accordance with the definitions for dermatologic immune-related adverse events defined by a recently completed international modified-Delphi consensus process for cirAE definitions[34]. We used trained dermatology reviewers to conduct a manual review of patient charts to phenotype these patients retrospectively for our study. Two independent dermatology reviewers categorized and graded suspected events using Common Terminology Criteria for Adverse Events version 5.0[11]. A score of 1 (highly unlikely) to 4 (highly likely) that these were cirAEs was assigned to each patient. Score was determined by manual review of patient charts and documentation which included images, morphology, histology, timing, competing medications, course of the rash, and response to treatment. We have added supplemental table 20, which outlines out of 676 patients diagnosed with cirAE, 218 (32.2%) had photographs, 134 (19.8%) had skin biopsies, and 372 (55%) were seen by a dermatologist within 2 years after ICI initiation. We added supplemental eFigure 2 and eFigure 3, which show timing of patients evaluated by a dermatologist within 2 years after ICI initiation and within 2 years after cirAE date respectively. A highly likely score was assigned if there was clinician documentation for immune-checkpoint inhibitor-related etiology, appropriate timing and morphology as defined in the above modified-Delphi consensus statement on cirAE definitions and prior literature on cirAEs, no competing medications that would contribute to development of cutaneous eruptions, and/or supportive evidence via photographs, histology, dermatologist or oncodermatologist evaluation. A likely score was assigned if there were competing medications present, but the competing medication is not likely to be the trigger given the timing or morphology of cirAE in relation to the competing medication. An unlikely score was assigned if given the timing of the ICI, there was a competing trigger more likely to have caused the eruption in comparison to ICI. A highly unlikely score was assigned if the timing of the cutaneous eruption did not coincide with timing of ICI or another cause for the cutaneous eruption was favored by the dermatologist. Patients with likelihood scores of 3 and 4 were categorized as having a cirAE. In cases that did not meet concordance between the two reviewers, the study's principal investigator, a board-certified dermatologist with expertise in cirAEs arbitrated the case. The concordance among the two independent reviewers prior to study PI arbitration was 88%. The calculated Copens kappa statistic or inter-rater reliability was 0.71. Based on the Cohen kappa statistic, we have substantial agreement for inter-rater reliability[35].

#### eFigures: 1

**eFigure 1:** Association of Vitiligo and Non-Vitiligo cirAEs with Survival Among ICI Recipients
**eFigure 2**: Timing of Dermatology Consultation After ICI Initiation Among Patients with CirAE Diagnosis
**eFigure 3:** Timing of Dermatology Consultation After cirAE Diagnosis

#### eTables: 19

**eTable 1:** Association of CirAE Status with Overall Survival Stratified by Cancer Type

**eTable 2**: Association of CirAE with Overall Survival Using a Multivariate Time-Varying Cox Proportional Hazards Model for Melanoma Cancer Patients
**eTable 3**: Association of CirAE with Overall Survival Using a Multivariate Time-Varying Cox Proportional Hazards Model in Non-melanoma Patients
**eTable 4.** Landmark Survival Analyses for Association of CirAE Development with Overall Survival in Patients Treated with ICI
**eTable 5:** Landmark Survival Analysis at 3 Months for Assocation of CirAE Development with Overall Survival in All Cancer Patients Treated with ICI
**eTable 6**: Landmark Survival Analysis at 6 Months for Association of CirAE Development with Overall Survival in All Cancer Patients Treated with ICI
**eTable 7**: Landmark Survival Analysis at 9 Months for Association of CirAE Development with Overall Survival in All Cancer Patients Treated with ICI
**eTable 8**: Landmark Survival Analysis at 1 Year for Association of CirAE Development with Overall Survival in All Cancer Patients Treated with ICI
**eTable 9**: Landmark Survival Analysis at 3 Months for Association of CirAE Development with Overall Survival in Melanoma Patients Treated with ICI
**eTable 10**: Landmark Survival Analysis at 6 Months for Association of CirAE Development with Overall Survival in Melanoma Patients Treated with ICI
**eTable 11**: Landmark Survival Analysis at 9 Months for Association of CirAE Development with Overall Survival in Melanoma Patients Treated with ICI
**eTable 12:** Landmark Survival Analysis at 1 Year for Association of CirAE Development with Overall Survival in Melanoma Patients Treated with ICI
**eTable 13**: Association of Vitiligo with Overall Survival Using a Multivariate Time-Varying Cox Proportional Hazards Model
**eTable 14**: Association of Acneiform Eruption with Overall Survival Using a Multivariate Time-Varying Cox Proportional Hazards Model
**eTable 15.** Association of Non-specific Rash with Overall Survival Using a Multivariate Time-Varying Cox Proportional Hazards Model
**eTable 16.** Association of Eczematous Eruption with Overall Survival Using a Multivariate Time-Varying Cox Proportional Hazards Model
**eTable 17.** Association of Isolated Pruritus (without visible manifestation of rash) with Overall Survival Using a Multivariate Time-Varying Cox Proportional Hazards Model
**eTable 18.** Association of Lichenoid Eruption with Overall Survival Using a Multivariate Time-Varying Cox Proportional Hazards Model
**eTable 19.** Association of Psoriasiform Eruption with Overall Survival Using a Multivariate Time-Varying Cox Proportional Hazards Model
**eTable 20.** Photographic, Skin Biopsy, and Dermatology Evaluation of Patients with CirAE Diagnosis

_**eFigure 1.** Association of Vitiligo and Non-Vitiligo cirAEs with Survival Among ICI Recipients

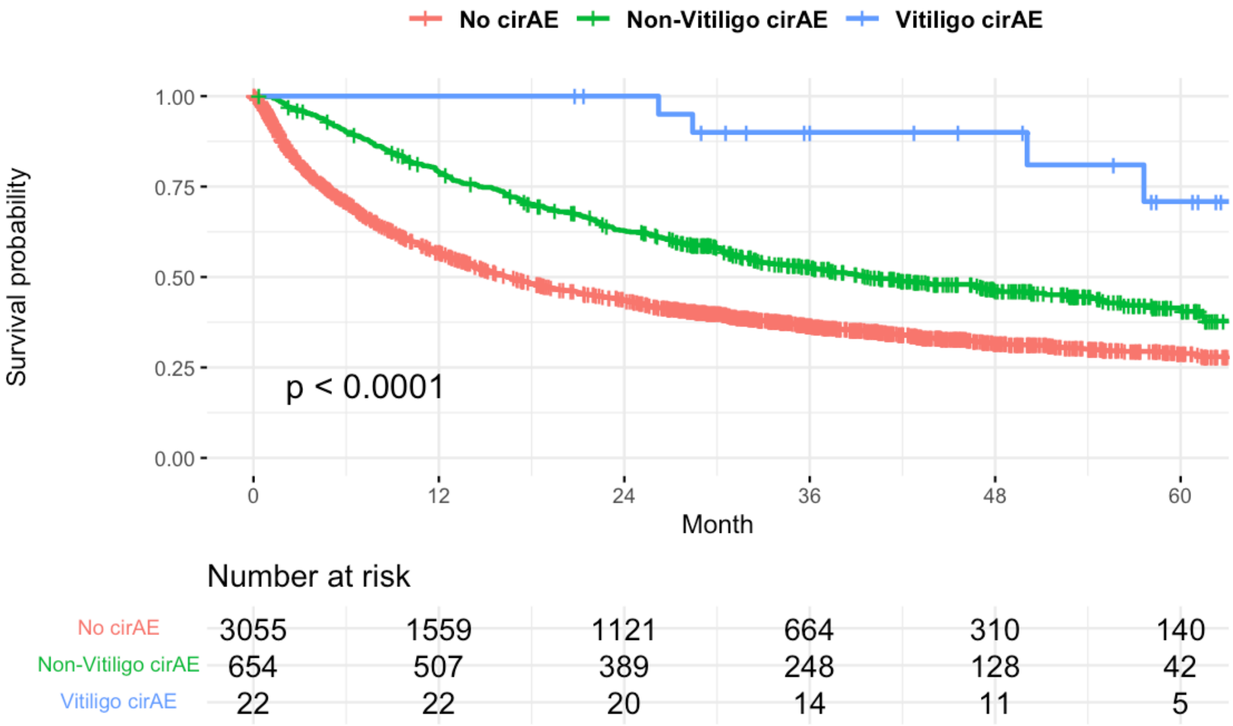


eFigure 1a. Association of cirAEs with overall survival among all ICI recipients. Comparison between no cirAE and non-vitiligo cirAE p-value<0.0001. Comparison between no cirAE and vitiligo p-value<0.0001.

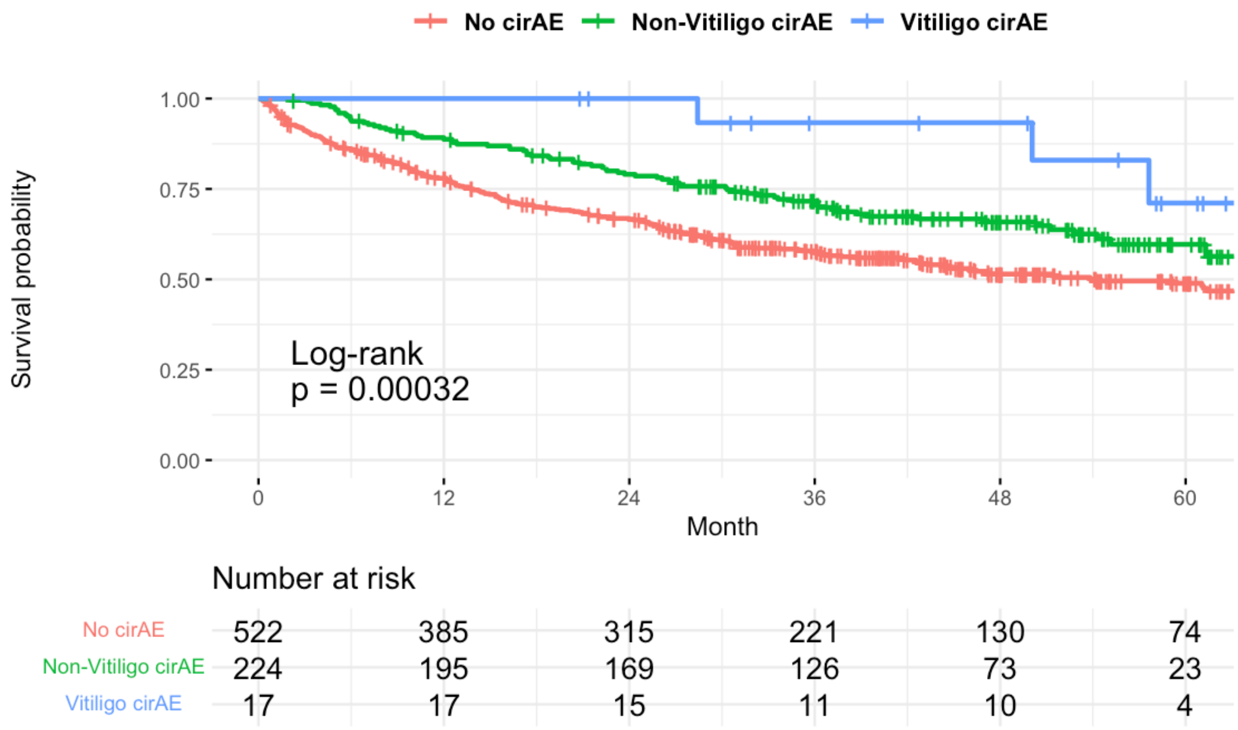


eFigure 1b. Association of cirAEs with overall survival among melanoma patients. Comparison between no cirAE and non-vitiligo cirAE p-value= 0.00072. Comparison between no cirAE and vitiligo cirAE p-value=0.02.

**eTable 1:** Association of CirAE with Overall Survival Stratified by Cancer Type

| **Cancer Type[a]** | **Hazard Ratio[b]** | **95% Confidence Interval** | **P-value** |
|---|---|---|---|
| Gynecologic | 0.62 | 0.32, 1.19 | 0.2 |
| Melanoma | 0.67 | 0.51, 0.87 | 0.003 |
| Sarcoma | 0.69 | 0.23, 2.03 | 0.5 |
| Head and Neck | 0.81 | 0.53, 1.23 | 0.3 |
| Gastrointestinal | 0.85 | 0.61, 1.17 | 0.3 |
| Thoracic | 0.96 | 0.75, 1.22 | 0.7 |
| Hematologic | 0.98 | 0.40, 2.39 | >0.9 |
| Genitourinary | 1.03 | 0.74, 1.43 | 0.9 |
| Non-melanoma Skin Cancer | 0.95 | 0.25, 3.56 | >0.9 |
| Breast | 1.39 | 0.79, 2.44 | 0.3 |
| Brain/Nervous | 1.43 | 0.74, 2.78 | 0.3 |

[a]Each row corresponds to a separate multivariable time-varying Cox PH model restricted to individual cancer type for the association of cirAE development with overall survival, adjusting for age at ICI initiation, sex, race, CCI, cancer type, ICI type, and year of ICI initiation.
[b]Hazard Ratio of the association of the presence of cirAE with overall survival.

**eTable 2**: Association of CirAE with Overall Survival Using a Multivariate Time-Varying Cox Proportional Hazards Model for Melanoma Cancer Patients

| Characteristic | Hazard Ratio | 95% Confidence Interval | P-value |
|---|---|---|---|
| **CirAE** | 0.67 | 0.51, 0.87 | 0.003 |
| **ICI Type** | | | |
| CTLA-4 | Ref[a] | Ref | |
| Combination Therapy | 1.74 | 1.12, 2.70 | 0.014 |
| PD-1 | 0.64 | 0.42, 0.97 | 0.037 |
| PD-L1 | 0.70 | 0.24, 2.02 | 0.5 |
| **Age of ICI Initiation** | 1.01 | 1.00, 1.03 | 0.014 |
| **Charlson Comorbidity Index** | 1.19 | 1.12, 1.27 | <0.001 |
| **Race/Ethnicity** | | | |
| White | Ref | Ref | |
| Asian | 0.71 | 0.18, 2.88 | 0.6 |
| Black | | | |
| Hispanic | 1.42 | 0.20, 10.2 | 0.7 |
| Other/Unknown | 0.30 | 0.04, 2.18 | 0.2 |
| **Sex** | | | |
| Female | Ref | Ref | |
| Male | 0.74 | 0.59, 0.93 | 0.011 |
| **Year of ICI Initiation** | 1.16 | 1.06, 1.26 | <0.001 |

[a]Ref= Reference group

**eTable 3**: Association of CirAE with Overall Survival Using a Multivariate Time-Varying Cox Proportional Hazards Model in Non-melanoma Patients

| Characteristic | Hazard Ratio | 95% Confidence Interval | P-value |
|---|---|---|---|
| **CirAE** | 0.92 | 0.81, 1.05 | 0.2 |
| **Charlson Comorbidity Index** | 1.08 | 1.06, 1.11 | <0.001 |
| **Age at ICI Initiation** | 0.99 | 0.99, 1.00 | 0.028 |
| **Sex** | | | |
| Female | Ref[a] | Ref | |
| Male | 1.08 | 0.98, 1.18 | 0.11 |
| **Race/Ethnicity** | | | |
| White | Ref | Ref | |
| Asian | 1.08 | 0.85, 1.36 | 0.5 |
| Black | 0.91 | 0.70, 1.18 | 0.5 |
| Hispanic | 0.98 | 0.57, 1.69 | >0.9 |
| Other/Unknown | 0.86 | 0.63, 1.17 | 0.3 |
| **ICI Type** | | | |
| CTLA-4 | Ref | Ref | |
| Combination Therapy | 1.04 | 0.63, 1.73 | 0.9 |
| PD-1 | 0.97 | 0.60, 1.56 | 0.9 |
| PD-L1 | 0.79 | 0.48, 1.30 | 0.3 |
| **Cancer Type** | | | |
| Thoracic | Ref | Ref | |
| Gastrointestinal | 1.32 | 1.15, 1.52 | <0.001 |
| Genitourinary | 0.79 | 0.68, 0.92 | 0.003 |
| Other | 0.91 | 0.82, 1.02 | 0.12 |
| **Year of ICI Initiation** | 1.04 | 0.99, 1.08 | 0.12 |

[a]Ref= Reference group

**eTable 4.** Landmark Survival Analyses for Association of CirAE Development with Overall Survival in Patients Treated with ICI

| Cancer Type | Landmark Time[a] | Number of Patients | Hazard Ratio[b] | 95% Confidence Interval | P-value |
|---|---|---|---|---|---|
| All Cancer | 3 months | 3059 | 0.75 | 0.67, 0.86 | <0.001 |
| | 6 months | 2648 | 0.83 | 0.72, 0.95 | 0.006 |
| | 9 months | 2335 | 0.89 | 0.77, 1.03 | 0.11 |
| | 12 months | 2092 | 0.94 | 0.80, 1.11 | 0.5 |
| Melanoma | 3 months | 711 | 0.65 | 0.50, 0.85 | 0.002 |
| | 6 months | 671 | 0.65 | 0.49, 0.87 | 0.003 |
| | 9 months | 633 | 0.64 | 0.47, 0.87 | 0.004 |
| | 12 months | 599 | 0.68 | 0.49, 0.94 | 0.021 |

[a]Each row corresponds to a separate multivariable Cox PH model, adjusting for age at ICI initiation, sex, race, CCI, cancer type, ICI type, and year of ICI initiation.
[b]Hazard Ratio of the assocation of the presence of cirAE wtih overall survival.

**eTable 5:** Landmark Survival Analysis at 3 Months for Association of CirAE Development with Overall Survival in All Cancer Patients Treated with ICI

| Characteristic | Hazard Ratio | 95% Confidence Interval | P-value |
|---|---|---|---|
| **CirAE** | 0.75 | 0.67, 0.85 | <0.001 |
| **Charlson Comorbidity Index** | 1.07 | 1.04, 1.09 | <0.001 |
| **Age of ICI Initiation** | 1.00 | 1.00, 1.01 | 0.6 |
| **Sex** | | | |
| Female | Ref[a] | Ref | |
| Male | 0.98 | 0.89, 1.09 | 0.7 |
| **Race/Ethnicity** | | | |
| White | Ref | Ref | |
| Asian | 0.94 | 0.70, 1.25 | 0.7 |
| Black | 0.89 | 0.65, 1.23 | 0.5 |
| Hispanic | 1.33 | 0.75, 2.36 | 0.3 |
| Other/Unknown | 0.75 | 0.51, 1.09 | 0.13 |
| **ICI Type** | | | |
| CTLA-4 | Ref | Ref | |
| Combination Therapy | 1.15 | 0.81, 1.64 | 0.4 |
| PD-1 | 0.87 | 0.63, 1.21 | 0.4 |
| PD-L1 | 0.77 | 0.54, 1.11 | 0.2 |
| **Cancer Type** | | | |
| Thoracic | Ref | Ref | |
| Gastrointestinal | 1.44 | 1.22, 1.70 | <0.001 |
| Genitourinary | 0.87 | 0.73, 1.03 | 0.11 |
| Melanoma | 0.51 | 0.43, 0.60 | <0.001 |
| Other | 1.04 | 0.91, 1.18 | 0.5 |
| **Year of ICI Initiation** | 1.06 | 1.01, 1.11 | 0.013 |

[a]Ref= Reference group

**eTable 6**: Landmark Survival Analysis at 6 Months for Association of CirAE Development with Overall Survival in All Cancer Patients Treated with ICI

| **Characteristic** | **Hazard Ratio** | **95% Confidence Interval** | **P-value** |
|---|---|---|---|
| **CirAE** | 0.83 | 0.72, 0.95 | 0.006 |
| **Charlson Comorbidity Index** | 1.05 | 1.02, 1.09 | <0.001 |
| **Age at ICI Initiation** | 1.00 | 1.00, 1.01 | 0.14 |
| **Sex** | | | |
| Female | Ref[a] | Ref | |
| Male | 1.00 | 0.89, 1.12 | >0.9 |
| **Race/Ethnicity** | | | |
| White | Ref | Ref | |
| Asian | 1.08 | 0.80, 1.47 | 0.6 |
| Black | 0.92 | 0.64, 1.32 | 0.6 |
| Hispanic | 1.44 | 0.74, 2.79 | 0.3 |
| Other/Unknown | 0.73 | 0.47, 1.13 | 0.2 |
| **ICI Type** | | | |
| CTLA-4 | Ref | Ref | |
| Combination Therapy | 1.03 | 0.71, 1.49 | 0.9 |
| PD-1 | 0.75 | 0.54, 1.06 | 0.10 |
| PD-L1 | 0.70 | 0.48, 1.02 | 0.061 |
| **Cancer Type** | | | |
| Thoracic | Ref | Ref | |
| Gastrointestinal | 1.38 | 1.14, 1.68 | 0.001 |
| Genitourinary | 0.86 | 0.70, 1.04 | 0.12 |
| Melanoma | 0.50 | 0.42, 0.60 | <0.001 |
| Other | 1.00 | 0.86, 1.15 | >0.9 |
| **Year of ICI Initiation** | 1.06 | 1.01, 1.12 | 0.016 |

[a]Ref= Reference group

**eTable 7**: Landmark Survival Analysis at 9 Months for Association of CirAE Development with Overall Survival in All Cancer Patients Treated with ICI

| Characteristic | Hazard Ratio | 95% Confidence Interval | P-value |
|---|---|---|---|
| **CirAE** | 0.89 | 0.77, 1.03 | 0.11 |
| **Charlson Comorbidity Index** | 1.06 | 1.02, 1.09 | 0.002 |
| **Age at ICI Initiation** | 1.01 | 1.00, 1.01 | 0.093 |
| **Sex** | | | |
| Female | Ref[a] | Ref | |
| Male | 0.94 | 0.83, 1.07 | 0.4 |
| **Race/Ethnicity** | | | |
| White | Ref | Ref | |
| Asian | 0.98 | 0.68, 1.41 | >0.9 |
| Black | 1.01 | 0.68, 1.50 | >0.9 |
| Hispanic | 1.47 | 0.70, 3.12 | 0.3 |
| Other/Unknown | 0.73 | 0.44, 1.19 | 0.2 |
| **ICI Type** | | | |
| CTLA-4 | Ref | Ref | |
| Combination Therapy | 0.89 | 0.60, 1.32 | 0.6 |
| PD-1 | 0.68 | 0.48, 0.96 | 0.029 |
| PD-L1 | 0.58 | 0.39, 0.87 | 0.008 |
| **Cancer Type** | | | |
| Thoracic | Ref | Ref | |
| Gastrointestinal | 1.33 | 1.06, 1.67 | 0.014 |
| Genitourinary | 0.93 | 0.75, 1.16 | 0.5 |
| Melanoma | 0.53 | 0.44, 0.64 | <0.001 |
| Other | 1.01 | 0.86, 1.20 | 0.9 |
| **Year of ICI Initiation** | 1.04 | 0.98, 1.10 | 0.2 |

[a]Ref= Reference group

**eTable 8**: Landmark Survival Analysis at 1 Year for Association of CirAE Development with Overall Survival in All Cancer Patients Treated with ICI

| Characteristic | Hazard Ratio | 95% Confidence Interval | P-value |
|---|---|---|---|
| **CirAE** | 0.94 | 0.80, 1.11 | 0.5 |
| **Charlson Comorbidity Index** | 1.05 | 1.01, 1.09 | 0.013 |
| **Age at ICI Initiation** | 1.01 | 1.00, 1.01 | 0.051 |
| **Sex** | | | |
| Female | Ref[a] | Ref | |
| Male | 0.95 | 0.82, 1.10 | 0.5 |
| **Race/Ethnicity** | | | |
| White | Ref | Ref | |
| Asian | 0.92 | 0.60, 1.41 | 0.7 |
| Black | 0.98 | 0.62, 1.55 | >0.9 |
| Hispanic | 1.74 | 0.77, 3.93 | 0.2 |
| Other/Unknown | 0.64 | 0.35, 1.17 | 0.15 |
| **ICI Type** | | | |
| CTLA-4 | Ref | Ref | |
| Combination Therapy | 1.06 | 0.68, 1.66 | 0.8 |
| PD-1 | 0.76 | 0.51, 1.14 | 0.2 |
| PD-L1 | 0.67 | 0.42, 1.05 | 0.083 |
| **Cancer Type** | | | |
| Thoracic | Ref | Ref | |
| Gastrointestinal | 1.16 | 0.89, 1.52 | 0.3 |
| Genitourinary | 0.90 | 0.70, 1.15 | 0.4 |
| Melanoma | 0.55 | 0.45, 0.68 | <0.001 |
| Other | 1.02 | 0.84, 1.22 | 0.9 |
| **Year of ICI Initiation** | 1.06 | 0.99, 1.13 | 0.084 |

[a]Ref= Reference group

**eTable 9**: Landmark Survival Analysis at 3 Months for Association of CirAE Development with Overall Survival in Melanoma Patients Treated with ICI

| Characteristic | Hazard Ratio | 95% Confidence Interval | P-value |
|---|---|---|---|
| **CirAE** | 0.65 | 0.50, 0.85 | 0.002 |
| **Charlson Comorbidity Index** | 1.14 | 1.06, 1.23 | <0.001 |
| **Age at ICI Initiation** | 1.02 | 1.00, 1.03 | 0.012 |
| **Sex** | | | |
| Female | Ref[a] | Ref | |
| Male | 0.74 | 0.58, 0.95 | 0.017 |
| **Race/Ethnicity** | | | |
| White | Ref | Ref | |
| Asian | 0.80 | 0.20, 3.23 | 0.8 |
| Black | | | |
| Hispanic | 1.57 | 0.22, 11.4 | 0.7 |
| Other/Unknown | 0.33 | 0.05, 2.38 | 0.3 |
| **ICI Type** | | | |
| CTLA-4 | Ref | Ref | |
| Combination Therapy | 1.55 | 0.97, 2.47 | 0.066 |
| PD-1 | 0.64 | 0.42, 0.99 | 0.047 |
| PD-L1 | 0.78 | 0.27, 2.7 | 0.6 |
| **Year of ICI Initiation** | 1.16 | 1.06, 1.27 | 0.002 |

[a]Ref= Reference group

**eTable 10**: Landmark Survival Analysis at 6 Months for Association of CirAE Development with Overall Survival in Melanoma Patients Treated with ICI

| Characteristic | Hazard Ratio | 95% Confidence Interval | P-value |
|---|---|---|---|
| **CirAE** | 0.65 | 0.49, 0.87 | 0.003 |
| **Charlson Comorbidity Index** | 1.11 | 1.02, 1.21 | 0.017 |
| **Age of ICI Initiation** | 1.02 | 1.01, 1.03 | 0.006 |
| **Sex** | | | |
| Female | Ref[a] | Ref | |
| Male | 0.76 | 0.58, 0.99 | 0.039 |
| **Race/Ethnicity** | | | |
| White | Ref | Ref | |
| Asian | 0.92 | 0.23, 3.73 | >0.9 |
| Black | | | |
| Hispanic | 1.96 | 0.27, 14.2 | 0.5 |
| Other/Unknown | 0.39 | 0.05, 2.79 | 0.3 |
| **ICI Type** | | | |
| CTLA-4 | Ref | Ref | |
| Combination Therapy | 1.53 | 0.94, 2.49 | 0.09 |
| PD-1 | 0.62 | 0.39, 0.98 | 0.039 |
| PD-L1 | 0.42 | 0.10, 1.81 | 0.2 |
| **Year of ICI Initiation** | 1.14 | 1.04, 1.26 | 0.007 |

[a]Ref= Reference group

**eTable 11**: Landmark Survival Analysis at 9 Months for Association of CirAE Development with Overall Survival in Melanoma Patients Treated with ICI

| Characteristic | Hazard Ratio | 95% Confidence Interval | P-value |
|---|---|---|---|
| **CirAE** | 0.64 | 0.47, 0.87 | 0.004 |
| **Charlson Comorbidity Index** | 1.13 | 1.03, 1.23 | 0.008 |
| **Age at ICI Initiation** | 1.02 | 1.00, 1.03 | 0.016 |
| **Sex** | | | |
| Female | Ref[a] | Ref | |
| Male | 0.66 | 0.50, 0.88 | 0.004 |
| **Race/Ethnicity** | | | |
| White | Ref | Ref | |
| Asian | 1.01 | 0.25, 4.11 | >0.9 |
| Black | | | |
| Hispanic | 2.41 | 0.33, 17.5 | 0.4 |
| Other/Unknown | 0.41 | 0.06, 2.98 | 0.4 |
| **ICI Type** | | | |
| CTLA-4 | Ref | Ref | |
| Combination Therapy | 1.41 | 0.85, 2.32 | 0.2 |
| PD-1 | 0.55 | 0.35, 0.87 | 0.011 |
| PD-L1 | 0.45 | 0.11, 1.94 | 0.3 |
| **Year of ICI Initiation** | 1.11 | 1.00, 1.23 | 0.050 |

[a]Ref= Reference group

**eTable 12:** Landmark Survival Analysis at 1 Year for Association of CirAE Development with Overall Survival in Melanoma Patients Treated with ICI

| **Characteristic** | **Hazard Ratio** | **95% Confidence Interval** | **P-value** |
|---|---|---|---|
| **CirAE** | 0.68 | 0.49, 0.94 | 0.021 |
| **Charlson Comorbidity Index** | 1.11 | 1.01, 1.23 | 0.040 |
| **Age of ICI Initiation** | 1.02 | 1.00, 1.04 | 0.009 |
| **Sex** | | | |
| Female | Ref[a] | Ref | |
| Male | 0.66 | 0.49, 0.89 | 0.006 |
| **Race/Ethnicity** | | | |
| White | Ref | Ref | |
| Asian | 1.15 | 0.28, 4.71 | 0.8 |
| Black | | | |
| Hispanic | 3.28 | 0.45, 23.9 | 0.2 |
| Other/Unknown | 0 | 0.00, Inf | >0.9 |
| **ICI Type** | | | |
| CTLA-4 | Ref | Ref | |
| Combination Therapy | 1.52 | 0.88, 2.63 | 0.13 |
| PD-1 | 0.58 | 0.35, 0.95 | 0.032 |
| PD-L1 | 0.53 | 0.12, 2.29 | 0.4 |
| **Year of ICI Initiation** | 1.11 | 1.00, 1.25 | 0.061 |

[a]Ref= Reference group

**eTable 13**: Association of Vitiligo with Overall Survival Using a Multivariate Time-Varying Cox Proportional Hazards Model

| Characteristic | Hazard Ratio | 95% Confidence Interval | P-value |
|---|---|---|---|
| **No CirAE** | Ref[a] | Ref | |
| **Non-vitiligo CirAE** | 0.70 | 0.64, 0.77 | <0.001 |
| **Vitiligo** | 0.29 | 0.12, 0.71 | 0.007 |
| **ICI Type** | | | |
| CTLA-4 | Ref | Ref | |
| Combination Therapy | 1.25 | 0.89, 1.72 | 0.2 |
| PD-1 | 0.96 | 0.71, 1.29 | 0.8 |
| PD-L1 | 0.82 | 0.59, 1.14 | 0.2 |
| **Charlson Comorbidity Index** | 1.09 | 1.06, 1.11 | <0.001 |
| **Age at ICI Initiation** | 1.00 | 0.99, 1.00 | 0.4 |
| **Race/Ethnicity** | | | |
| White | Ref | Ref | |
| Asian | 1.04 | 0.80, 1.35 | 0.8 |
| Black | 0.89 | 0.67, 1.18 | 0.4 |
| Hispanic | 0.93 | 0.50, 1.75 | 0.8 |
| Other/Unknown | 0.78 | 0.55, 1.10 | 0.2 |
| **Sex** | | | |
| Female | Ref | Ref | |
| Male | 1.03 | 0.94, 1.13 | 0.6 |
| **Cancer Type** | | | |
| Thoracic | Ref | Ref | |
| Gastrointestinal | 1.32 | 1.13, 1.53 | <0.001 |
| Genitourinary | 0.79 | 0.67, 0.92 | 0.003 |
| Melanoma | 0.43 | 0.37, 0.50 | <0.001 |
| Other | 0.93 | 0.83, 1.04 | 0.2 |
| **Year of ICI Initiation** | 1.05 | 1.01, 1.10 | 0.011 |

[a]Ref= Reference group

**eTable 14**: Association of Acneiform Eruption with Overall Survival Using a Multivariate Time-Varying Cox Proportional Hazards Model

| Characteristic | Hazard Ratio | 95% Confidence Interval | P-value |
|---|---|---|---|
| **No CirAE** | Ref[a] | Ref | |
| **Non-acneiform eruption CirAE** | 0.70 | 0.64, 0.76 | <0.001 |
| **Acneiform eruption** | 0.34 | 0.13, 0.87 | 0.025 |
| **ICI Type** | | | |
| CTLA-4 | Ref | Ref | |
| Combination Therapy | 1.23 | 0.89, 1.71 | 0.2 |
| PD-1 | 0.95 | 0.71, 1.29 | 0.8 |
| PD-L1 | 0.82 | 0.59, 1.13 | 0.2 |
| **Charlson Comorbidity Index** | 1.09 | 1.06, 1.11 | <0.001 |
| **Age at ICI Initiation** | 1.00 | 0.99, 1.00 | 0.4 |
| **Race/Ethnicity** | | | |
| White | Ref | Ref | |
| Asian | 1.04 | 0.80, 1.35 | 0.7 |
| Black | 0.87 | 0.66, 1.16 | 0.4 |
| Hispanic | 0.91 | 0.48, 1.71 | 0.8 |
| Other/Unknown | 0.78 | 0.55, 1.10 | 0.2 |
| **Sex** | | | |
| Female | Ref | Ref | |
| Male | 1.03 | 0.94, 1.13 | 0.5 |
| **Cancer Type** | | | |
| Thoracic | Ref | Ref | |
| Gastrointestinal | 1.32 | 1.13, 1.54 | <0.001 |
| Genitourinary | 0.79 | 0.67, 0.92 | 0.003 |
| Melanoma | 0.43 | 0.37, 0.49 | <0.001 |
| Other | 0.93 | 0.83, 1.05 | 0.2 |
| **Year of ICI Initiation** | 1.05 | 1.01, 1.10 | 0.010 |

[a]Ref= Reference group

**eTable 15.** Association of Non-specific Rash with Overall Survival Using a Multivariate Time-Varying Cox Proportional Hazards Model

| Characteristic | Hazard Ratio | 95% Confidence Interval | P-value |
|---|---|---|---|
| **No CirAE** | Ref[a] | Ref | |
| **Non-rash NOS CirAE** | 0.69 | 0.62, 0.78 | <0.001 |
| **Rash NOS** | 0.68 | 0.57, 0.81 | <0.001 |
| **ICI Type** | | | |
| CTLA-4 | Ref | Ref | |
| Combination Therapy | 1.23 | 0.89, 1.71 | 0.2 |
| PD-1 | 0.95 | 0.71, 1.29 | 0.8 |
| PD-L1 | 0.82 | 0.59, 1.13 | 0.2 |
| **Charlson Comorbidity Index** | 1.09 | 1.06, 1.11 | <0.001 |
| **Age at ICI Initiation** | 1.00 | 0.99, 1.00 | 0.4 |
| **Race/Ethnicity** | | | |
| White | Ref | Ref | |
| Asian | 1.04 | 0.80, 1.35 | 0.8 |
| Black | 0.88 | 0.66, 1.16 | 0.4 |
| Hispanic | 0.91 | 0.49, 1.71 | 0.8 |
| Other/Unknown | 0.78 | 0.56, 1.11 | 0.2 |
| **Sex** | | | |
| Female | Ref | Ref | |
| Male | 1.03 | 0.94, 1.13 | 0.6 |
| **Cancer Type** | | | |
| Thoracic | Ref | Ref | |
| Gastrointestinal | 1.32 | 1.13, 1.54 | <0.001 |
| Genitourinary | 0.79 | 0.67, 0.92 | 0.003 |
| Melanoma | 0.43 | 0.37, 0.49 | <0.001 |
| Other | 0.93 | 0.82, 1.04 | 0.2 |
| **Year of ICI Initiation** | 1.05 | 1.01, 1.10 | 0.010 |

[a]Ref= Reference group

**eTable 16.** Association of Eczematous Eruption with Overall Survival Using a Multivariate Time-Varying Cox Proportional Hazards Model

| Characteristic | Hazard Ratio | 95% Confidence Interval | P-value |
|---|---|---|---|
| **No CirAE** | Ref[a] | Ref | |
| **Non-eczematous Eruption CirAE** | 0.69 | 0.63, 0.75 | <0.001 |
| **Eczematous Eruption** | 0.71 | 0.50, 1.01 | 0.056 |
| **ICI Type** | | | |
| CTLA-4 | Ref | Ref | |
| Combination Therapy | 1.23 | 0.89, 1.71 | 0.2 |
| PD-1 | 0.95 | 0.71, 1.29 | 0.8 |
| PD-L1 | 0.82 | 0.59, 1.13 | 0.2 |
| **Charlson Comorbidity Index** | 1.09 | 1.06, 1.11 | <0.001 |
| **Age at ICI Initiation** | 1.00 | 0.99, 1.00 | 0.4 |
| **Race/Ethnicity** | | | |
| White | Ref | Ref | |
| Asian | 1.04 | 0.80, 1.35 | 0.8 |
| Black | 0.88 | 0.66, 1.16 | 0.4 |
| Hispanic | 0.91 | 0.49, 1.71 | 0.8 |
| Other/Unknown | 0.78 | 0.56, 1.11 | 0.2 |
| **Sex** | | | |
| Female | Ref | Ref | |
| Male | 1.03 | 0.94, 1.13 | 0.6 |
| **Cancer Type** | | | |
| Thoracic | Ref | Ref | |
| Gastrointestinal | 1.32 | 1.13, 1.54 | <0.001 |
| Genitourinary | 0.79 | 0.67, 0.92 | 0.003 |
| Melanoma | 0.43 | 0.37, 0.49 | <0.001 |
| Other | 0.93 | 0.82, 1.04 | 0.2 |
| **Year of ICI Initiation** | 1.05 | 1.01, 1.10 | 0.010 |

[a]Ref= Reference group

**eTable 17.** Association of Isolated Pruritus (without visible manifestation of rash) with Overall Survival Using a Multivariate Time-Varying Cox Proportional Hazards Model

| Characteristic | Hazard Ratio | 95% Confidence Interval | P-value |
|---|---|---|---|
| **No CirAE** | Ref[a] | Ref | |
| **Non-isolated Pruritus CirAE** | 0.69 | 0.62, 0.76 | <0.001 |
| **Isolated Pruritus** | 0.71 | 0.55, 0.91 | 0.007 |
| **ICI Type** | | | |
| CTLA-4 | Ref | Ref | |
| Combination Therapy | 1.23 | 0.89, 1.71 | 0.2 |
| PD-1 | 0.95 | 0.71, 1.29 | 0.8 |
| PD-L1 | 0.82 | 0.59, 1.13 | 0.2 |
| **Charlson Comorbidity Index** | 1.09 | 1.06, 1.11 | <0.001 |
| **Age at ICI Initiation** | 1.00 | 0.99, 1.00 | 0.5 |
| **Race/Ethnicity** | | | |
| White | Ref | Ref | |
| Asian | 1.04 | 0.80, 1.35 | 0.8 |
| Black | 0.88 | 0.66, 1.16 | 0.4 |
| Hispanic | 0.91 | 0.48, 1.71 | 0.8 |
| Other/Unknown | 0.78 | 0.56, 1.11 | 0.2 |
| **Sex** | | | |
| Female | Ref | Ref | |
| Male | 1.03 | 0.94, 1.13 | 0.6 |
| **Cancer Type** | | | |
| Thoracic | Ref | Ref | |
| Gastrointestinal | 1.32 | 1.13, 1.54 | <0.001 |
| Genitourinary | 0.79 | 0.67, 0.92 | 0.003 |
| Melanoma | 0.43 | 0.37, 0.49 | <0.001 |
| Other | 0.93 | 0.82, 1.04 | 0.2 |
| **Year of ICI Initiation** | 1.05 | 1.01, 1.10 | 0.010 |

**eTable 18.** Association of Lichenoid Eruption with Overall Survival Using a Multivariate Time-Varying Cox Proportional Hazards Model

| Characteristic | Hazard Ratio | 95% Confidence Interval | P-value |
|---|---|---|---|
| **No CirAE** | Ref[a] | Ref | |
| **Non-lichenoid Eruption CirAE** | 0.71 | 0.65, 0.78 | <0.001 |
| **Lichenoid Eruption** | 0.51 | 0.36, 0.73 | <0.001 |
| **ICI Type** | | | |
| CTLA-4 | Ref | Ref | |
| Combination Therapy | 1.24 | 0.89, 1.72 | 0.2 |
| PD-1 | 0.96 | 0.71, 1.29 | 0.8 |
| PD-L1 | 0.82 | 0.59, 1.14 | 0.2 |
| **Charlson Comorbidity Index** | 1.09 | 1.06, 1.11 | <0.001 |
| **Age at ICI Initiation** | 1.00 | 0.99, 1.00 | 0.5 |
| **Race/Ethnicity** | | | |
| White | Ref | Ref | |
| Asian | 1.04 | 0.80, 1.35 | 0.8 |
| Black | 0.88 | 0.66, 1.16 | 0.4 |
| Hispanic | 0.91 | 0.48, 1.71 | 0.8 |
| Other/Unknown | 0.78 | 0.55, 1.10 | 0.2 |
| **Sex** | | | |
| Female | Ref | Ref | |
| Male | 1.03 | 0.94, 1.13 | 0.6 |
| **Cancer Type** | | | |
| Thoracic | Ref | Ref | |
| Gastrointestinal | 1.31 | 1.13, 1.53 | <0.001 |
| Genitourinary | 0.79 | 0.67, 0.92 | 0.003 |
| Melanoma | 0.43 | 0.37, 0.49 | <0.001 |
| Other | 0.93 | 0.83, 1.04 | 0.2 |
| **Year of ICI Initiation** | 1.05 | 1.01, 1.10 | 0.011 |

[a]Ref= Reference group

**eTable 19.** Association of Psoriasiform Eruption with Overall Survival Using a Multivariate Time-Varying Cox Proportional Hazards Model

| Characteristic | Hazard Ratio | 95% Confidence Interval | P-value |
|---|---|---|---|

| | | | |
|---|---|---|---|
| **No CirAE** | Ref[a] | Ref | |
| **Non-psoriasiform Eruption CirAE** | 0.70 | 0.64, 0.76 | <0.001 |
| **Psoriasiform Eruption** | 0.52 | 0.33, 0.82 | 0.005 |
| **ICI Type** | | | |
| CTLA-4 | Ref | Ref | |
| Combination Therapy | 1.23 | 0.89, 1.71 | 0.2 |
| PD-1 | 0.96 | 0.71, 1.29 | 0.8 |
| PD-L1 | 0.82 | 0.59, 1.14 | 0.2 |
| **Charlson Comorbidity Index** | 1.09 | 1.06, 1.11 | <0.001 |
| **Age at ICI Initiation** | 1.00 | 0.99, 1.00 | 0.5 |
| **Race/Ethnicity** | | | |
| White | Ref | Ref | |
| Asian | 1.04 | 0.80, 1.35 | 0.8 |
| Black | 0.88 | 0.66, 1.17 | 0.4 |
| Hispanic | 0.91 | 0.49, 1.71 | 0.8 |
| Other/Unknown | 0.79 | 0.56, 1.11 | 0.2 |
| **Sex** | | | |
| Female | Ref | Ref | |
| Male | 1.03 | 0.94, 1.13 | 0.5 |
| **Cancer Type** | | | |
| Thoracic | Ref | Ref | |
| Gastrointestinal | 1.32 | 1.13, 1.54 | <0.001 |
| Genitourinary | 0.79 | 0.67, 0.92 | 0.003 |
| Melanoma | 0.43 | 0.37, 0.49 | <0.001 |
| Other | 0.93 | 0.83, 1.04 | 0.2 |
| **Year of ICI Initiation** | 1.05 | 1.01, 1.10 | 0.01 |

[a]Ref= Reference group

**eTable 20:** Photographic, Skin Biopsy, and Dermatology Evaluation of Patients with CirAE Diagnosis

| | | **Total N=676** |
|---|---|---|
| Photograph | No | 458 (67.8%) |

| | | |
|---|---|---|
| | Yes | 218 (32.2%) |
| Skin Biopsy | No | 542 (80.2%) |
| | Yes | 134 (19.8%) |
| Dermatology Evaluation | No | 304 (45.0%) |
| | Yes | 372 (55.0%) |
